# Strategy to step on obstacle during cross-country race to minimize overuse injury risk

**DOI:** 10.1101/2025.10.08.25337622

**Authors:** Emanuela H. Kang, David G. Wilson, Jan K. Petric, Sophia Ulman

**Affiliations:** Hockaday School, Dallas, Texas; Department of Prosthetics-Orthotics, University of Texas Southwestern Medical Center, Dallas, Texas; Orthopaedic and Sports Medicine Center, Scottish Rite for Children, Frisco, Texas; Department of Orthopaedic Surgery, University of Texas Southwestern Medical Center, Dallas, Texas

**Keywords:** medial-stepping, in-silico, computational model, overuse injury, safe running

## Abstract

Overuse injuries are a major concern in high school endurance sports such as cross-country running. This study explores how joint laxity, foot placement, and surface friction affect ligament strain during obstacle interaction, using an in-silico computer model simulating a “stepping on obstacle” scenario. Incorporating ground reaction forces and muscle activation, the model evaluates soft tissue responses across 27 cases—varying three levels each of joint laxity (none, medium, high), foot placement (central, medial, lateral), and surface friction (low, medium, high). Ligament strain was recorded at every millisecond and analyzed statistically. Results show foot placement is the dominant factor in minimizing ligament strain, with medial foot strikes yielding the lowest anterior tibiotalar ligament strain (11.5%), which increased by 1.75× and 1.73× in central and lateral strikes, respectively. Friction level showed a negative correlation with strain, reducing anterior tibiotalar stress by 17% from low to high friction. Joint laxity positively correlated with plantar fascia strain, with the medium laxity group experiencing 1.17× more strain than the no-laxity group. These findings suggest that medial foot placement and higher surface friction significantly reduce injury risk, providing biomechanical insight and practical guidance for safer running techniques in young endurance athletes.

## Introduction

Overuse injuries present a significant challenge in high school athletics, particularly in endurance sports like cross-country running [1-3]. Among high school cross-country runners, overuse injuries impact approximately 28-38% of athletes within a single season [3]. These injuries are commonly associated with repetitive stress and inadequate recovery [1-3], which are inherent to the sport due to high training volumes and intensities. The impact from overuse injuries is substantial, as they often require extensive medical rehabilitation and physical therapy that can last for months [1,3]. This extended recovery period places a considerable burden on athletes, coaches, and healthcare systems. Additionally, these injuries can negatively affect performance, disrupt season continuity, and pose long-term health risks to young athletes.

Overuse injuries are especially concerning in cross-country runners because, unlike acute injuries, these athletes often continue training despite experiencing pain. This continued activity magnifies the damage, leading to cumulative microtrauma that can progressively weaken bone and connective tissues [1,3,4]. Furthermore, research shows that the speed of ligament recovery is much slower compared to bone [4-7]. Once damaged, ligaments often don’t heal completely due to persistent stress [5,6], leading to chronic pain, reduced performance, and a higher risk of re-injury [3,4,7]. Continuous use of unhealed ligaments compromise athletes’ long-term performance and health leading to long-term complications. Therefore, taking preventive measures to avoid injury to tissues, especially ligaments, are crucial for athletes to maintain their health and athletic abilities over time.

A key step towards solving this problem is finding ways to improve running techniques, particularly to protect individual tissues, and analyze what factors have the most significant impact on these tissues. For this reason, this study takes an in-silico model approach similar to other sports movement studies [8-12]. In-silico models, which are studies performed on a computer or via computer simulation, have been widely used in understanding real-life questions especially in the engineering field [4,9,13]. With evolution of computational processing capability and support of biomaterial study, biomechanical studies have actively used in-silico models [11,12,14-17]. The model can simulate various scenarios by independently controlling each factor [13,18] and allows for the examination of the stress on the bones and strain on soft tissues non-invasively and simultaneously [8,16,12,18]. The optimal combination of the factors among various running scenarios would be identified as well as the relative influence of each factor. In addition, extreme conditions [16,18,19], which may lead to critical injury, can be tested without ethical issues. By identifying these influences on individual tissues, evidence-based strategies [14,15,17] can be developed to help runners minimize stress and strain, further reduce the risk of injury, and ultimately guide safer running practices.

The purpose of this study is to investigate how different factors influence ligament strain during running over obstacles. There are two objectives: (1) to evaluate the impact of each factor—joint laxity, foot placement, and surface friction—on the strain experienced by ligaments in the foot, and (2) to provide recommendations for safer running techniques that reduce the likelihood of ligament injury. This research aims to fill the gap in knowledge with data-driven insights that can guide runners toward safer practices while encountering obstacles. The study is based on three key hypotheses that aim to identify the safest running conditions for athletes: (1) the original level of joint laxity, representing a state where the legs are not fatigued, is the safest condition; (2) the central foot placement, specifically at the center of the plantar surface of the foot, provides the safest positioning; and (3) high friction surfaces are hypothesized to be safer, compared to low friction surfaces. These hypotheses are designed to test various factors that could influence the safety of runners, guiding future recommendations for injury prevention.

## Materials and methods

An in-silico computer model was developed to simulate a "stepping on obstacle" scenario during running. This model aimed to investigate the behavior of tissues in the foot when faced with an external challenge, such as stepping on uneven surfaces, common during cross-country running. By using an in-silico approach, the ethical concerns associated with human subjects, particularly those involving injury risk, can be taken away. The in-silico model approach was chosen since it allows for the analysis of internal forces and motions inside the body and for assessing factors independently and repeatedly under controlled conditions.

For this study, a custom shoe model was developed and integrated into an existing foot in-silico model (Fig 1). The foot model is part of the THUMS model [16], which was initially developed for injury analysis in the automotive industry. The THUMS model is widely recognized and utilized by not only automotive manufacturers but also by insurance companies, clinical researchers, and governmental agencies including the National Highway Safety Administration and the Food and Drug Administration [8,9,12,18,19]. The model is freely available, has been extensively validated for its representativeness in simulating human biomechanics [10,14-16,18], and has been used to analyze the biomechanics in diverse human populations [9,10,12]. The foot model itself used in this study has already been validated [10,16] and includes 26 bones (26 cortical; 26 trabecular),18 ligaments, 1 tendon, and other soft tissues. In addition, the shoe model includes the shoe outsole, shoe bottom, and the shoe top. The impactor and ground plates are introduced to replicate the obstacle on the ground. The shoe model is developed using a preprocessing tool (Hypermesh, Altair Engineering, Inc., Troy, MI) and the simulation is performed using analysis tool (LS-DYNA, ANSYS, Inc., Canonsburg, PA).

**Fig 1.**
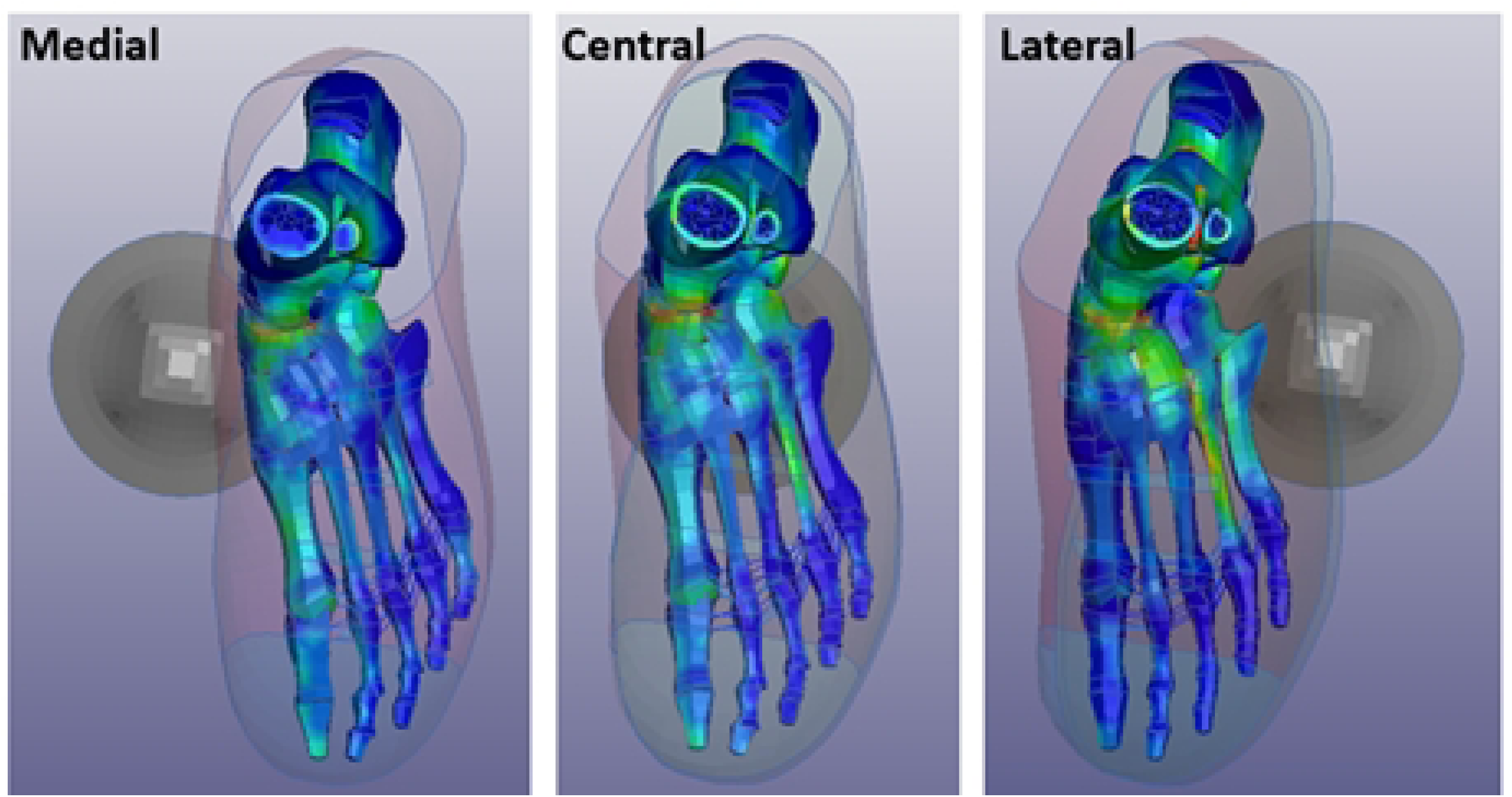
Schematic images of the stepping on obstacle in-silico model setup.

In this analysis, 47,218 volumetric elements and 16,130 shell elements were used to represent the human foot-ankle-knee and the interacting obstacles on the ground. The skin (all except joint area), cortical bones, tendons, ligaments, and membranes were expressed as thin shell elements having four nodes. Other soft tissues including fat and muscle and trabecular bones were modeled as hexahedral elements. The skin in the joint area was expressed as triangular shell and soft tissues in the joint were modeled as tetrahedron models in order to represent the geometrical features to connect between knee and foot. The average mesh size is about 0.5mm and mesh sensitivity study has been already performed by THUMS model itself, so the mesh sizes for others were also maintained as the same mesh size to reduce the unnecessary contact definition.

To simulate the running motion in a computationally efficient way, ground reaction forces were applied to the foot by moving the obstacle upward to impact the foot from below, while the proximal ends of the tibia and fibula were fixed. The mean peak-to-peak force component of the ground reaction force during the running cycle in vertical direction is applied on the impactor [20]. The ground plate stayed fixed throughout the simulation, with only the impactor moving to replicate the forces of 3 times the body weight encountered at the foot during a running cycle [20]. In addition, the model accounted for the change in velocity of the center of gravity during the support phase, set at 3.15 ms^-1^. This ensured that the simulation closely mimicked real world running biomechanics [20].

According to the ground reaction forces analysis study in distance running, the midfoot consistently experiences the highest impact during the running cycle, regardless of the first contact location [20]. Thus, the midfoot served as the impactor point in the simulation. To understand the effect of impact location, the obstacle is positioned to interact with the foot at three different locations: the medial, central, and lateral regions of the foot (Fig 2).

**Fig 2.**
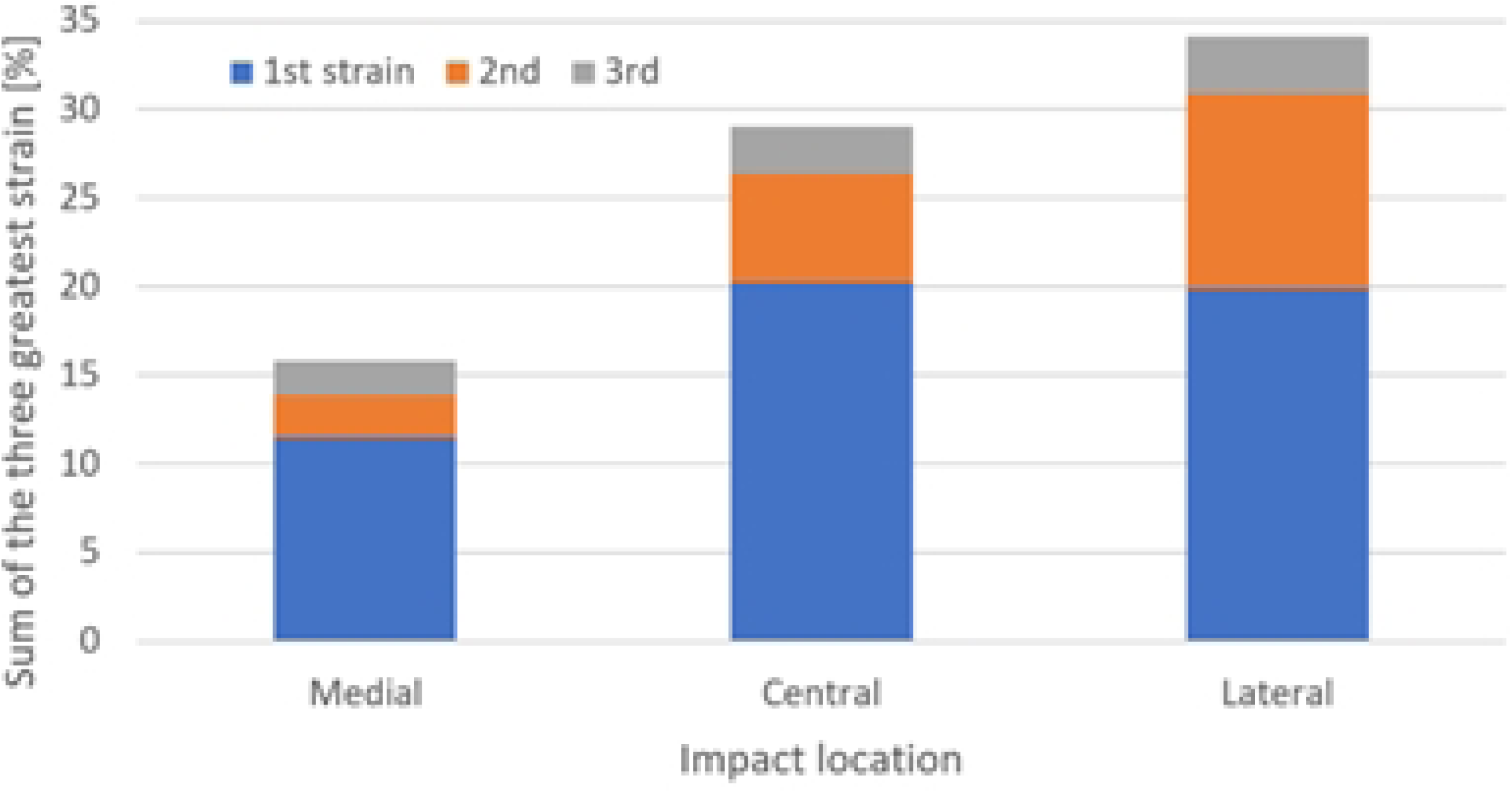
Three impacting locations (Medial, Central, Lateral) in the mid-foot.

To define the contact interactions within the simulation, three key interfaces were established: impactor vs shoe, shoe vs foot, and ground vs shoe. The friction coefficients at contact points were varied to simulate different running and shoe conditions at three levels: 0.2 (slippery), 0.5 (mild), and 0.8 (high friction), similar to other footwear design simulation studies. This variation allowed for a detailed analysis of how different friction levels affect foot stability and the sensitivity to the risk of injury during running.

The material properties of the shoe were assumed to provide adequate damping, ensuring realistic force absorption during impact [13,21]. Material properties (Young’s modulus and Poisson’s ratio) for the bones, tendon, ligaments, and shoe components used in the simulation are outlined in Table 1. The information of muscles, flesh, and skin are not provided in Table 1, because their more-complicated behavior could not be captured with few linear elastic material parameters. The detailed viscoelastic nonlinear behavior including muscle activation information could be found directly from the THUMS. On the other hand, the impactor and the ground plate are assumed as rigid, thus those are also not included in Table 1. To account for joint laxity, the stiffness of the ligaments surrounding the ankle joint was adjusted, simulating a reduction in their structural integrity [14,17] as research reported that joint laxity can increase by approximately 30% within one hour of exercise [4,15]. The joint laxity levels were expressed as three levels: no (original joint), medium (30%), and high (60%) laxity. These adjustments are accounted in the model to represent the biomechanical changes that occur during prolonged running.

**Table 1.**
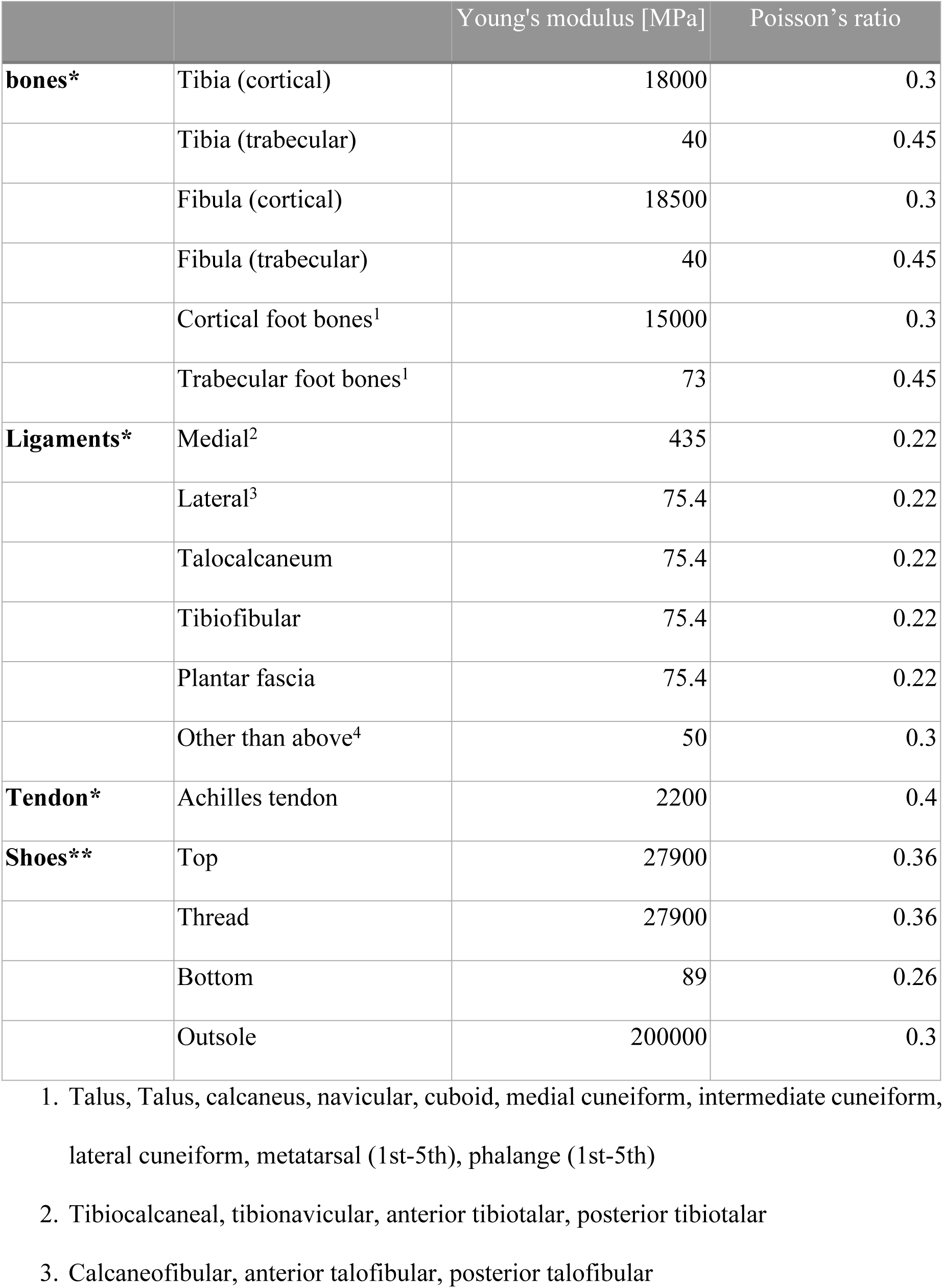
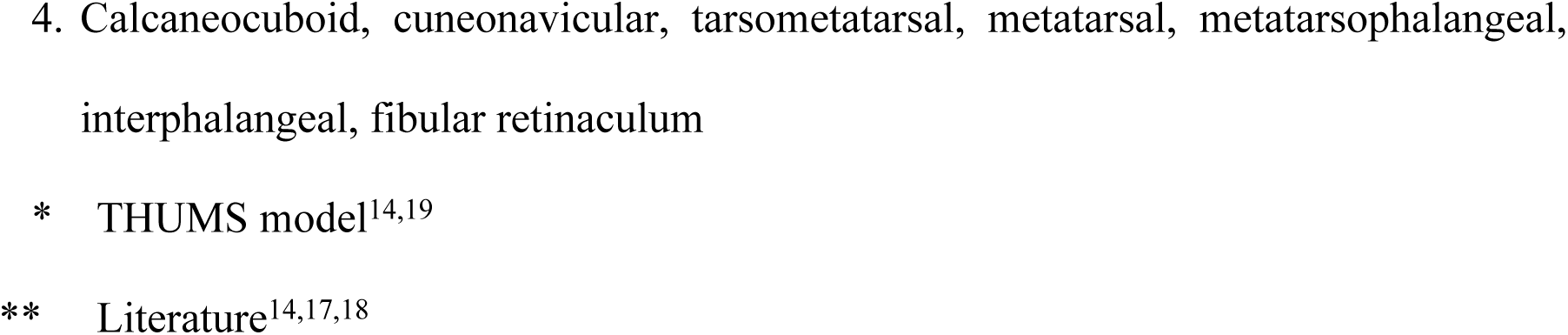
Material property information of key components in the model.

|  |  | Young's modulus [MPa] | Poisson's ratio |
| --- | --- | --- | --- |
| **bones\*** | Tibia (cortical) | 18000 | 0.3 |
|  | Tibia (trabecular) | 40 | 0.45 |
|  | Fibula (cortical) | 18500 | 0.3 |
|  | Fibula (trabecular) | 40 | 0.45 |
|  | Cortical foot bones<sup>1</sup> | 15000 | 0.3 |
|  | Trabecular foot bones<sup>1</sup> | 73 | 0.45 |
| **Ligaments\*** | Medial<sup>2</sup> | 435 | 0.22 |
|  | Lateral<sup>3</sup> | 75.4 | 0.22 |
|  | Talocalcaneum | 75.4 | 0.22 |
|  | Tibiofibular | 75.4 | 0.22 |
|  | Plantar fascia | 75.4 | 0.22 |
|  | Other than above<sup>4</sup> | 50 | 0.3 |
| **Tendon\*** | Achilles tendon | 2200 | 0.4 |
| **Shoes\*\*** | Top | 27900 | 0.36 |
|  | Thread | 27900 | 0.36 |
|  | Bottom | 89 | 0.26 |
|  | Outsole | 200000 | 0.3 |

1. Talus, Talus, calcaneus, navicular, cuboid, medial cuneiform, intermediate cuneiform, lateral cuneiform, metatarsal (1st-5th), phalange (1st-5th) 2. Tibiocalcaneal, tibionavicular, anterior tibiotalar, posterior tibiotalar 3. Calcaneofibular, anterior talofibular, posterior talofibular
4. Calcaneocuboid, cuneonavicular, tarsometatarsal, metatarsal, metatarsophalangeal, interphalangeal, fibular retinaculum
\* THUMS model<sup>14,19</sup>
\*\* Literature<sup>14,17,18</sup>

Throughout the 27 simulation cases (3 joint laxity levels x 3 foot placement locations x 3 friction levels), strain values in the ligaments were predicted and recorded at 1 msec intervals as the primary observation. The stress distribution in the bones was also collected for understanding the transferring pathway of the impact. The influence of each factor—joint laxity, impact location, and friction level— was analyzed using one-way ANOVA, followed by post-hoc paired t-tests to determine statistically significant differences between the conditions. Furthermore, regression analysis was performed to indicate the relative strength of individual factors. The analysis was performed using SPSS Statistics (IBM SPSS, Inc., Chicago, IL, USA). A significance level of p = 0.05 was applied to all statistical analyses.

## Results

In all of the 27 “stepping on obstacle” simulation scenarios (Table 2), the anterior tibiotalar ligament demonstrated the greatest strain (17.2±4.4%), followed by the plantar fascia (6.5±3.7%). The third largest strain values (2.6±1.0%) depended on the condition; tibiocalcaneal (with all lateral foot placement; with central strike in a loose joint with low or mild friction level), posterior tibiotalar (with medial strike in a loose joint, regardless of friction level; with central strike in a loose joint with high friction), or tibionavicular (in original joint condition with central or medial foot placement, regardless of friction level).

**Table 2.** Summary of strain value predicted in 27 cases.

| Joint condition | Impact location | Friction level | 1st greatest strain [%], ATTL | 2nd greatest strain [%], PFL | 3rd greatest strain [%], ligament |
| --- | --- | --- | --- | --- | --- |
| No laxity | **Central** | 0.2 | 21.42 | 5.10 | 2.19 |
|  |  | 0.5 | 20.26 | 5.16 | 1.39 |
|  |  | 0.8 | 18.41 | 8.84 | 1.52 |
|  | Lateral | 0.2 | 21.24 | 9.70 | 2.72 |
|  |  | 0.5 | 19.07 | 10.55 | 2.86 |
|  |  | 0.8 | 18.11 | 10.56 | 2.91 |
|  | Medial | 0.2 | 14.18 | 2.36 | 2.06 |
|  |  | 0.5 | 12.45 | 2.37 | 2.13 |
|  |  | 0.8 | 12.23 | 1.77 | 1.72 |
| Medium laxity | **Central** | 0.2 | 23.02 | 7.04 | 1.38 |
|  |  | 0.5 | 18.48 | 6.03 | 3.99 |
|  |  | 0.8 | 18.57 | 9.07 | 4.14 |
|  | Lateral | 0.2 | 21.18 | 12.65 | 3.27 |
|  |  | 0.5 | 18.48 | 11.61 | 3.46 |
|  |  | 0.8 | 18.02 | 12.05 | 3.41 |
|  | Medial | 0.2 | 12.56 | 2.02 | 1.53 |
|  |  | 0.5 | 11.52 | 2.77 | 2.25 |
|  |  | 0.8 | 11.19 | 3.00 | 2.20 |
| High laxity | Central | 0.2 | 23.50 | 4.95 | 4.02 |
|  |  | 0.5 | 20.48 | 4.66 | 4.28 |
|  |  | 0.8 | 17.52 | 4.70 | 1.01 |
|  | Lateral | 0.2 | 22.74 | 10.81 | 3.56 |
|  |  | 0.5 | 20.95 | 10.52 | 3.28 |
|  |  | 0.8 | 19.11 | 10.33 | 4.12 |
|  | Medial | 0.2 | 10.93 | 1.67 | 1.54 |
|  |  | 0.5 | 9.85 | 2.82 | 1.93 |
|  |  | 0.8 | 8.76 | 2.43 | 2.14 |

The greatest strain, observed at the anterior tibiotalar ligament (fourth column in Table 2), exhibited a strong relationship with foot placement (p = 1.3e-12, medial vs. central) and friction level (p = 1.8e-5), but not with joint condition. The regression model showed high predictive accuracy (R² = 0.938). First, the lowest anterior tibiotalar ligament strain was predicted in the medial foot placement scenarios (Fig 3). At the corresponding joint condition and friction levels, the peak strain values in the medial impact location were as small as 46-63% of the values in the central or lateral cases. The average strain for medial impact was predicted at 11.5%, increasing by 1.75 times in the central group (p = 1.6e-6) and 1.73 times in the lateral group (p = 9.0e-7). Second, significantly reduced anterior tibiotalar ligament was also observed with higher friction level (Fig 4). For example, the mean strain of 19.0% in the 0.2 friction group decreased by 0.89 times in the 0.5 friction group (p = 2.5e-4) and further to 0.83 times in the 0.8 friction group (p = 6.2e-5). The increased benefit in reducing the strain by changing friction level from low (0.2) to high (0.8) was observed in the higher joint laxity groups, although this reduction did not reach statistical significance (p=0.073 for medium-high laxity groups; 0.084 for no-high laxity groups). Third, increased joint laxity was associated with greater variability in anterior tibiotalar ligament strain values (Fig 3). The standard deviations of peak strain were 3.4, 4.0, and 5.4% for no, medium, and high laxity groups, respectively. Particularly in the central foot placement condition, the standard deviations were 1.2%, 2.1%, and 2.4% for the respective laxity levels. Although joint laxity was not a statistically significant predictor in the overall regression model, stratified analyses revealed its influence under specific foot placement and friction conditions. For example, with focusing on the worst foot placement case, the lateral impacting scenario, the average of the anterior tibiotalar ligament strain (19.5%) among the three friction levels in no laxity joint group increased to 20.9% (p=0.015) in high laxity groups. Similarly, with restricting the analysis to the loose joint condition (medium and high laxity groups), the average strain value in the lateral case was 1.86 times of the average strain in the medial case (p=6.3e-5).

**Fig 3.**
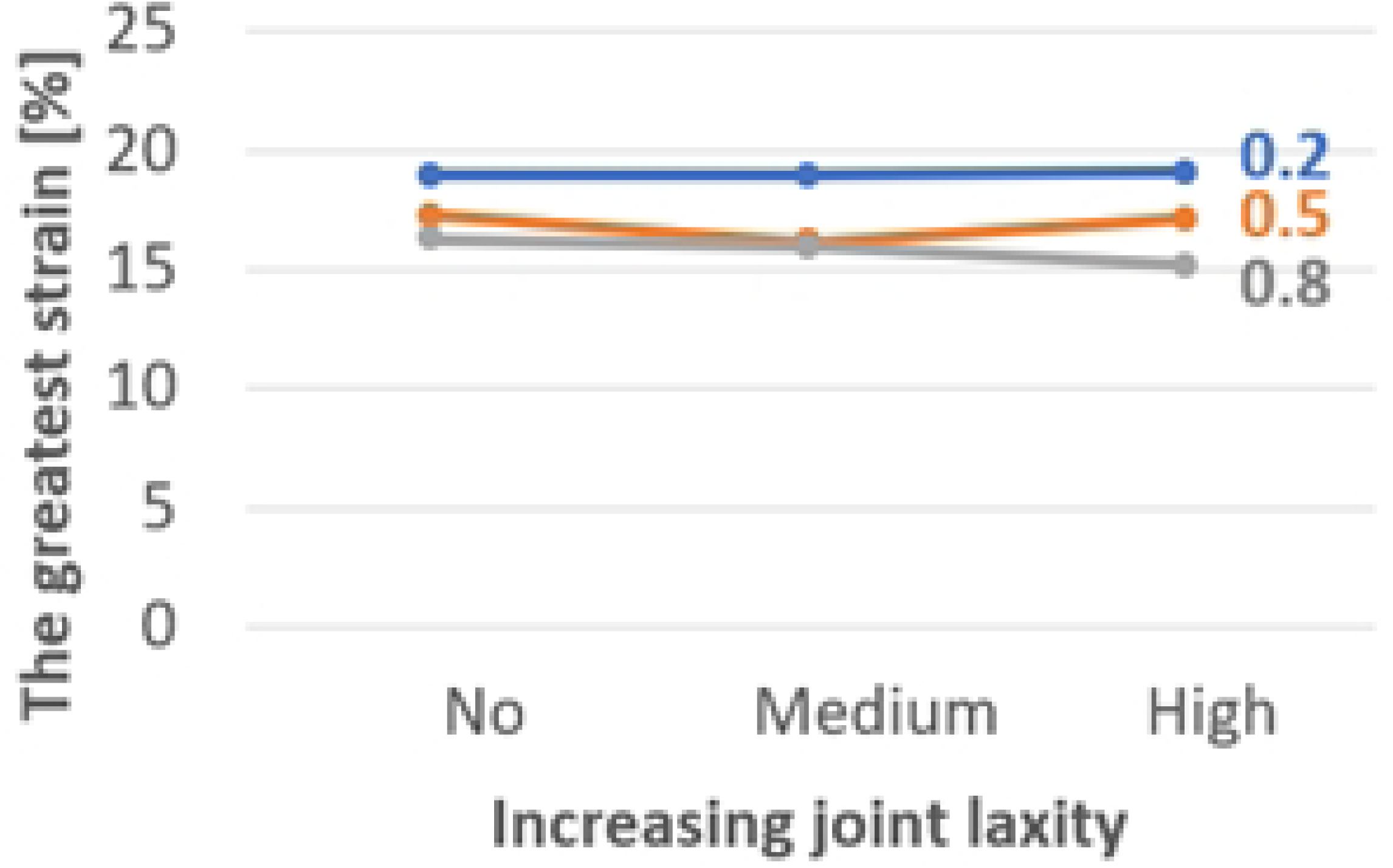
The mean and standard deviation of the strain on the anterior tibiotalar ligament in three impact locations.

**Fig 4.**
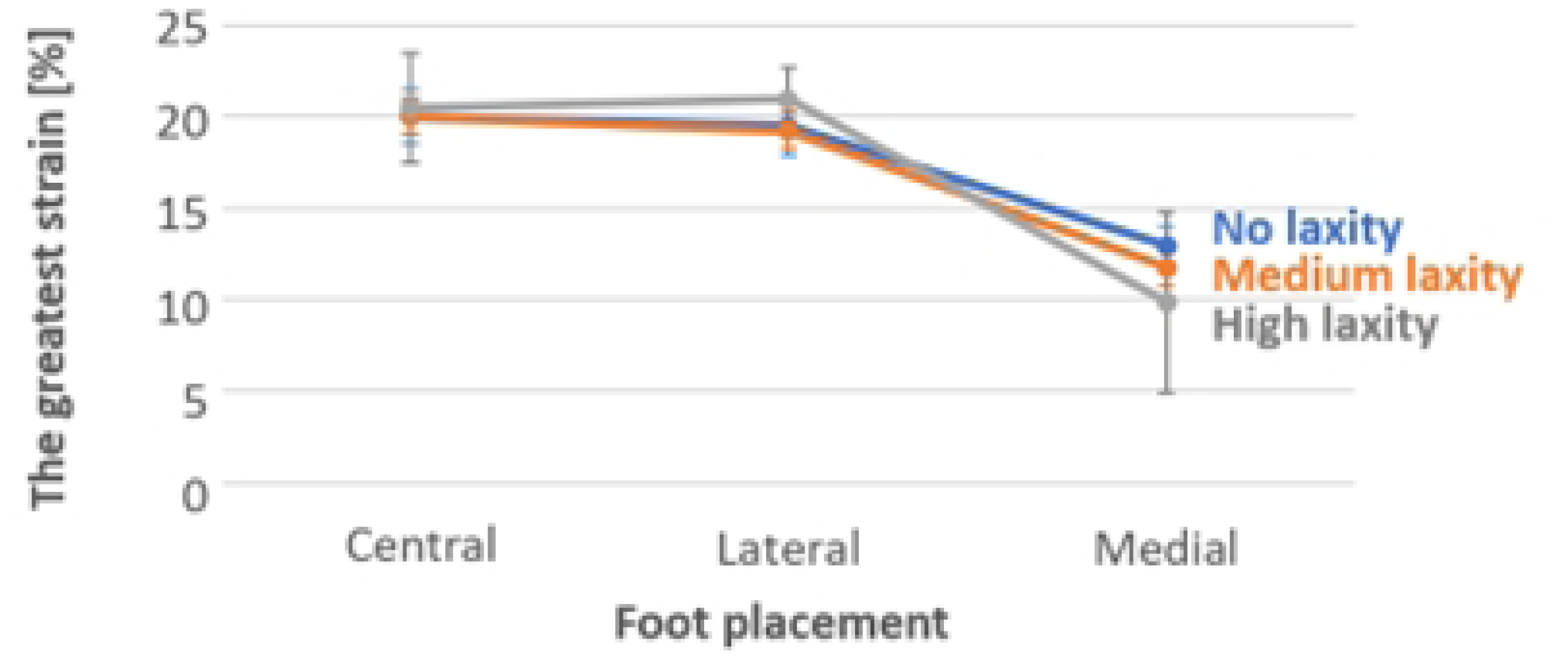
The average strain values among three impact location scenarios in each joint laxity condition and friction condition.

The second greatest strain values, predicted on the plantar fascia (fifth column in Table 2), showed a strong relationship with both foot placement (p = 6.9e-11) and joint condition (p = 0.0042), but not with friction level. The regression model yielded a high predictive power (R² = 0.945). First, significantly lower plantar fascia strain was predicted for the medial impact location compared to the other two impact locations (p = 5.0e-8 for medial vs. central; p = 1.3e-11 for medial vs. lateral). Lateral impact resulted in an average strain of approximately 11% (11.0 ± 1.6), while central impact led to about 6% strain (6.2 ± 0.9). In contrast, medial impact produced only about 2% strain (2.4 ± 0.4). Second, increased plantar fascia strain was also associated with greater joint laxity. For example, the average strain in the medium laxity group was 1.17 times higher than in the no-laxity group (p = 0.0051). Focusing specifically on lateral impact scenarios, the average strain (10.3%) in the no-laxity group increased by 1.18 times in the medium laxity group (p = 0.043). Third, no significant influence of friction level on plantar fascia strain was observed.

The third greatest strain value (the last column in Table 2) also showed an increasing tendency with both a more lateral impact location (medial to lateral; p=0.032) and higher joint laxity level (p=0.090), as indicated by a moderate predictive regression model (R^2^=0.424). First, the average third greatest strain in the medial impact group (1.9%) increased by 1.37 times in the central impact group (p = 0.083) and by 1.69 times in the lateral impact group (p = 1.79e-5). Second, the average value in the no-laxity group (2.17%) increased by 1.31 times in the medium laxity group (p = 0.064) and by 1.33 times in the high laxity group (p = 0.049). When focusing on lateral impact scenarios, the average third greatest strain (2.83%) in the no-laxity group increased by 1.19 times in the medium laxity group (p = 1.4e-3) and by 1.29 times in the high laxity group (p = 3.5e-2). Third, friction level was not observed to have any influence on the third greatest strain value.

Among the three factors, foot impact location had the greatest influence on strain values. This effect was illustrated in Fig 5, where the averages of the three highest strain values were summed for each foot placement group. Specifically, the anterior tibiotalar ligament strain, plantar fascia strain, and the third greatest strain, typically observed in one of the medial ligament complexes, were averaged across the three joint laxity levels and three friction conditions.

**Fig 5.**
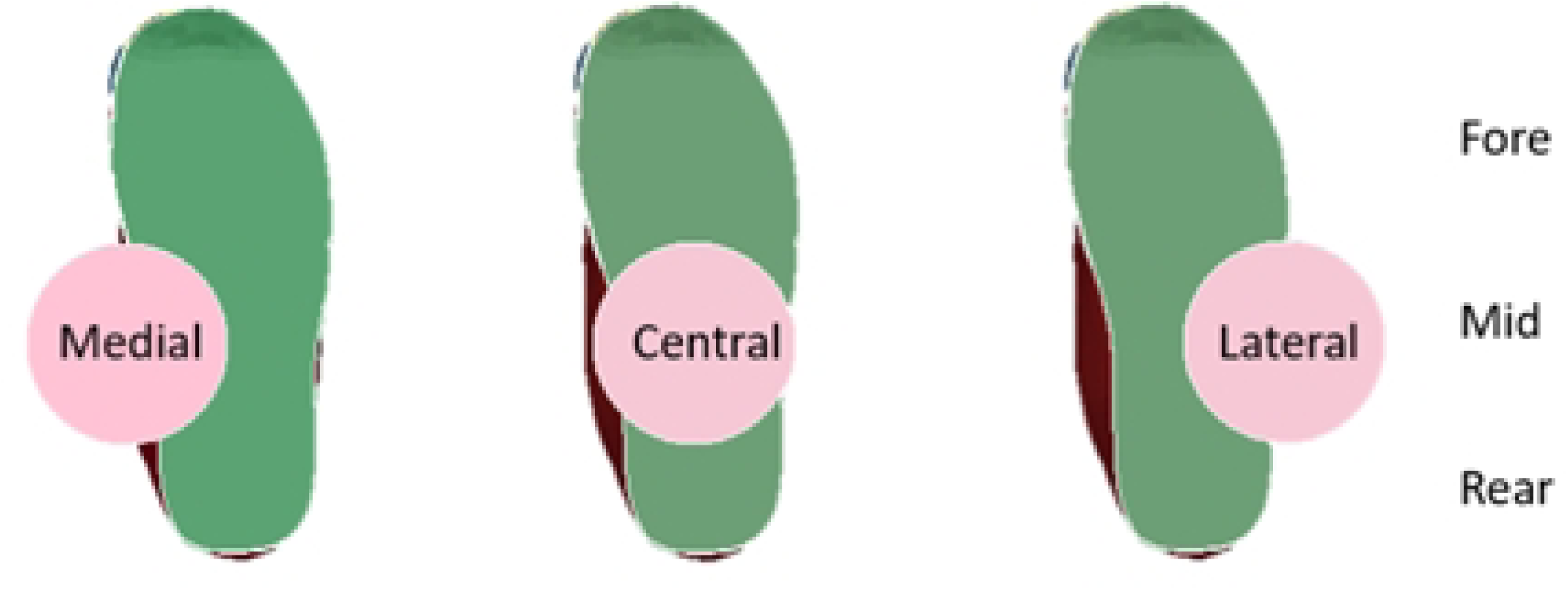
The sum of the three greatest strain average values in each foot placement group.

The stress distribution in the bones, which was collected and analyzed specifically to provide reference information on load transfer through the foot, was also influenced by the impact location (Fig 6). The results shown in Fig 6 represent the condition with high joint laxity and mild friction. Lateral impact resulted in a relatively focal stress distribution, with minimal stress dissipation through the big toe. In contrast, medial impact led to broader dispersion of stress through the arch, soft tissues, and hallux, in addition to the big toe. The stress pattern observed in the central impact case was intermediate, combining features of both the medial and lateral impact scenarios.

**Fig 6.**
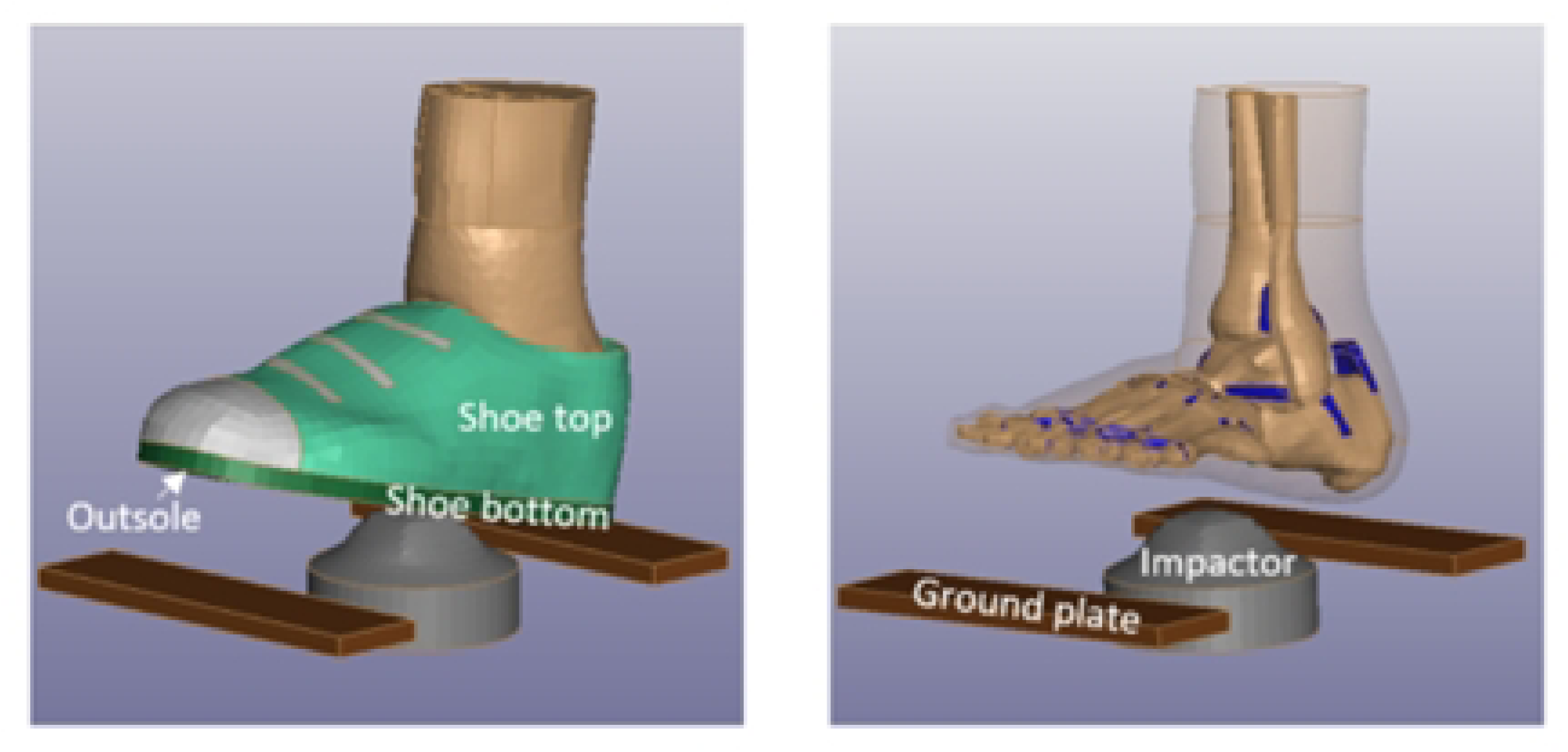
Stress distribution in the foot at three different impacting loading cases with 60% of joint laxity and 0.5 friction coefficient, at the impact moment.

## Discussion

This study aimed to identify the safest running condition when encountering obstacles by examining the effects of joint laxity, foot placement, and friction on the ligament strains. The findings highlight foot placement as the most influential factor, with minimal strain observed when obstacles were encountered on the medial side of the foot—supporting the rejection of the second hypothesis. Neither the first (joint laxity) nor third (friction) hypotheses could be statistically confirmed or rejected across all conditions. However, the first hypothesis appears to hold partial support in relation to the second and third highest strain magnitudes or when the confounding effect of foot placement is removed. Likewise, the third hypothesis finds statistical support when strain on the anterior tibiotalar ligament is considered in isolation.

Regarding the first hypothesis, a positive trend was observed between joint laxity level and ligament strain, however, a statistically significant relationship was found only for the plantar fascia. This may be due to the increased variability in strain values associated with higher joint laxity. Such variability could amplify or obscure the effects of other contributing factors. These findings are consistent with those of a previous in-silico walking simulation study [17], which examined the impact of generalized ligament laxity on the progression of hallux valgus deformity. Their simulations demonstrated that increased laxity broadened the range of peak stress and medial–lateral foot displacement. As a result, they highlighted the potential for either amplified or masked effects of hallux valgus during specific phases of gait (e.g., the first and/or second peaks of the stance phase). Furthermore, considering that joint laxity has been reported to increase by approximately 30% after one hour of running [4,15], taking regular breaks, ideally every hour, may help reduce strain accumulation in the foot ligaments, especially plantar fascia.

Regarding the second hypothesis, the results clearly demonstrated that ligaments experienced significantly lower peak strain under medial impact conditions compared to central and lateral impacts. The medial side of the foot forms the arch and plays a primary role in shock absorption and weight distribution. In addition, the distal end of the lower limb is anatomically aligned more medially rather than centrally relative to the foot, as illustrated in Fig 6. Therefore, stepping on the medial side of the foot promotes more efficient impact dissipation and weight distribution by aligning the direction of impact with the bone-supported axis of energy transfer. Furthermore, relatively large bones, such as the first metatarsal and proximal phalanges, are positioned on the medial side of the foot. When impact is transferred through this region, an effective dissipation pathway is supported by these robust structures. These biomechanical features help explain the significantly lower peak strain observed in the medial impact condition. The current results suggest that medial foot strikes may help mitigate strain on key soft tissue structures during running, potentially lowering the risk of injury.

As for the third hypothesis, higher friction was associated with a statistically significant reduction in anterior tibiotalar ligament strain, regardless of joint laxity condition. This finding suggests that increasing friction can help reduce ligament strain during running, thereby potentially lowering the risk of anterior tibiotalar ligament injury. Moreover, the strain-reducing effect of increased friction was more pronounced at higher joint laxity levels, with trends approaching statistical significance. This observation implies that higher friction may partially compensate for joint laxity by enhancing stability during foot-ground interaction.

Among the various ligaments examined, the medial ligament complex and the plantar fascia exhibited two of the top three greatest strain values. The medial ligament complex is a strong, flat, triangular structure that connects the tibia to the navicular, calcaneus, and talus. It plays a critical role in ankle joint stability and in transmitting forces between the tibia and the tarsal bones. This mechanical function may explain the strong relationship observed between strain in the medial ligament complex (first and third highest strain values) and both foot placement and joint condition. A significant negative relationship between anterior tibiotalar ligament strain and friction level suggests that higher friction may contribute to ankle joint stability by reducing mechanical demand on this ligament. Meanwhile, the plantar fascia—a thick band of connective tissue running from the heel to the toes along the sole of the foot—helps maintain arch integrity and absorb impact through both the arch and the fascia itself. Given this function, it is not surprising that both foot placement and joint laxity significantly influenced strain in the plantar fascia. Furthermore, once injured, ligaments such as these tend to heal slowly, and complete offloading of the affected structures during the healing process is often impractical [4,5]. Therefore, proactively minimizing strain and potential damage to these critical soft tissue structures should be prioritized as a preventive strategy [1,4,22].

Notably, all predicted strain values in this study remained within the microtrauma range. Failure strain has been reported in in-vitro tests using elderly cadaveric feet—which typically exhibit lower mechanical tolerance than in-vivo tissues or those from healthy young individuals—as ranging from 6% to 17% [4,7,23]. In comparison, failure strain in the human Achilles tendon has been measured at 44.3% and 49.2% under faster and slower displacement rates, respectively [7,24]. The peak strain values predicted in the present study reached approximately 20%, remaining well below the thresholds commonly associated with acute ligament failure. Therefore, while stepping on an obstacle is unlikely to result in immediate acute injury, it may still induce microtrauma. Over time, cumulative microtrauma can contribute to tissue degeneration and eventual failure [4,22].

This study is not without limitations. First, although the focus was on human biomechanics, the effects of shoe design and variation in midsole damping were not considered. These factors could influence the impact dissipation through the footwear and, consequently, modify the stress and strain experienced by the foot. Second, individual differences in foot morphology—such as soft tissue distribution, arch height, and toe alignment—were not included in the simulation model. These anatomical variations may influence the risk of overuse injury and should be explored in future research. Finally, incorporating these personalized factors to better understand and reduce foot stress and strain could support the development of individualized running strategies, which represents a long-term goal of this research direction.

## Implications

Statistically significant reductions in soft tissue strain were predicted in the medial foot placement scenario. Effective dissipation of impact in this condition was further supported by the observed stress distribution patterns. These findings strongly suggest that, when stepping on obstacles during endurance activities such as cross-country running, athletes should aim to contact the medial side of the foot. This strategy may help protect critical structures such as the medial ligament complex and plantar fascia from overuse injuries. In addition, increased joint laxity places a greater mechanical burden on these tissues. As previous studies have shown that joint laxity may increase after prolonged activity, taking breaks at regular intervals—ideally every hour—is recommended to reduce cumulative strain. Furthermore, this study highlights the importance of maintaining high friction at the foot-ground interface. Under fatigued conditions that elevate joint laxity, increased friction may help reduce ligament strain and mitigate the risk of overuse injuries that often require extended recovery periods.

## Conclusions

To reduce the risk of injury during running, particularly when stepping on obstacles, this study independently examined the effects of joint laxity, foot placement, and surface friction on ligament strain in the foot. The results demonstrated a significant reduction in strain when the foot contacted the obstacle on its medial side. This finding suggests that medial foot placement is the most effective strategy for minimizing overuse injury risk during obstacle negotiation in running. Additionally, under fatigued conditions that increase joint laxity, maintaining high friction at the foot-ground interface may help limit the associated rise in ligament strain. Implementing these strategies may help reduce the mechanical burden on critical foot structures and mitigate the risk of injuries that typically require long recovery periods.

## Data Availability

Data Availability Statement: All simulation files and datasets generated during the current study will be available from the database (DOI to be provided upon acceptance).

## Disclosure statement

The authors report no conflicts of interest or financial disclosures related to this study.

## Acknowledgements

We gratefully acknowledge Dr. Barbara Fishel, Dr. Jen Fore, and the Hockaday School Writing Center for mentorship, constructive feedback on the study design, and sustained support throughout this project.

## Notes

### Competing Interest Statement

The authors have declared no competing interest.

### Funding Statement

The author(s) received no specific funding for this work.

### Author Declarations

This study did not involve human subjects and therefore did not require approval or oversight from an Institutional Review Board (IRB) or equivalent body.

